# COVID-19: a study about the impact of coronavirus on physicians of La Plata, Argentina

**DOI:** 10.1101/2021.11.05.21265969

**Authors:** María Virginia Croce, Elsa Chiappa, Adriana Moiso, Martin E Rabassa

## Abstract

**Background:** In Argentina, the burden of COVID-19 on health systems and physicians was substantial with difficulties on daily triage decisions which have to be made in the context of grave shortages of basic equipment and consumables.

**Purpose:** this study was performed to understand what physicians were experiencing during the COVID-19 pandemic in La Plata (capital city of Buenos Aires province, Argentina).

**Methods:** A cross-sectional study was performed; a questionnaire was sent by e-mail to physicians who work in this city during November 2020. The questionnaire was made based on Medscape US and International Physicians’ COVID-19 Experience Report: Risk, Burnout, Loneliness.

**Statistical analysis:** test for normality was performed employing the Kolmogorov-Smirnov test while Chi-square test of independence to examine the relationship between sex and workplace with nominal variables. For categorical variables, Kendall’s tau correlation was performed to test for independence. ANOVA was developed to examine differences between physician’s age. Statistical significance was set to p < 0.05 in all cases. All statistical analysis was done employing SPSS Statistics, Version 24 (IBM, USA).

**Results:** 203 physicians answered the questionnaire; the majority of physicians (96%) considered stressful their experience during pandemic and reported distress episodes being for more than 60% the most stressful of their practices, 30% presented depression and were medically treated, while 32.7% felt loneliness with 4 physicians with suicidal thoughts.

**Conclusion:** The results highlight the need to protect the psychological well-being of the healthcare community, and to invest resources to significantly promote the mental health of professionals.

## Introduction

The World Health Organization (WHO) has declared a public health emergency of international concern over the global outbreak of COVID-19 on 30 January 2020, and has escalated it to a global pandemic on 11 March Coronavirus Disease (COVID-19) Outbreak (https://www.who.int.).

Physicians suffer mental and physical pressures during the COVID-19 pandemic; physicians, among health-care workers, provide care for patients despite exhaustion, personal risk of infection, fear of transmission to family members, illness or death of friends and colleagues, and the loss of many patients. They also face many additional sources of stress and anxiety, and long shifts combined with unprecedented population restrictions, including personal isolation, have affected their individuals’ physical and mental health^1^.

In Argentina, the burden of COVID-19 on health systems and health-care workers was substantial with difficulties on daily triage decisions which have to be made in the context of grave shortages of basic equipment and consumables. An increase in non-COVID-19-related health problems and deaths further strained a poorly resourced health system.

Objective: this study was performed to understand what physicians were experiencing during the COVID-19 pandemic in La Plata (capital city of Buenos Aires province, Argentina).

## Methods

A cross-sectional study was performed in La Plata, the capital city of Buenos Aires Province (Argentina); a questionnaire was sent by e-mail to physicians who work in this city (District I Medical College) during November 2020. The questionnaire was made based on Medscape US and International Physicians’ COVID-19 Experience Report: Risk, Burnout, Loneliness^2^. The research was approved by the Medical Bioethics Committee, Faculty of Medical Sciences, National University of La Plata, Argentina, reference N°101/21. Informed consent was obtained from all individual participants included in the study, data were anonymized to preserve confidentiality.

### Statistical analysis

Test for normality was performed by employing the Kolmogorov-Smirnov test while Chi-square test of independence to examine the relationship between sex and workplace with nominal variables. For categorical variables, Kendall’s tau correlation was performed to test for independence. ANOVA was employed to examine differences between physician’s age. Statistical significance was set to p < 0.05 in all cases. No power analysis was performed. All statistical analysis was done employing SPSS Statistics, Version 24 (IBM, USA).

## Results

A total of 203 answers were collected from physicians aged 28-72 years old, 122 women and 81 men; 43 (21.2%) worked at the Private Health System only, 39 (19.2%) at the Public System while 121 (59.6%) in both Health Systems. Clinical Medicine was the most frequent specialty found (64/203, 31.5%), followed by Intensive Care (17/203, 8.37%), Surgical specialties (15/203, 7.38%), Infectious diseases (14/203, 6.9%), and others. More than 90% physicians had treated COVID-19 patients; most of them personally (115/203, 56.65%), 53/203 (26.1%) personally and via video or phone, 17/203 (8.4%) via video or phone but not personally, and only 18/203 (8.9%) had not treated COVID-19 patients. Physicians working at the Public System were more likely to treat patient personally than physicians working at the Private System or at both systems, X^2^ (6, N=203) = 19.500, p=0.003. Also, surgeons treated COVID-19 patients exclusively in person, while clinicians opted for personally and video interviews (X^2^ (6, N = 203) = 20.955, p =0.002). Finally, ANOVA showed that mean age of physicians who treated patients personally was lower than those who use video assisted meetings (F (3,199) = 13.890, p < 0.001), Post hoc comparisons using the Tukey’s B test also indicated that the mean age of physicians who treat COVID-19 patients personally was lower (N=115, M=44.97 years) than video assisted interviews (N=17, M=57.76 years).

While 57/185 (30.81%) treated patients with COVID-19 without having the appropriate PPE (these answers included always, often, and sometimes), 12.9% (26/202) had been diagnosed with COVID-19. Forty-eight physicians (23.6%) said they had immediate family members who were diagnosed with COVID-19; it was unknown if the diagnosis was either confirmed by a test or symptoms. Men were more likely than women to report a family member with COVID-19 diagnosis. The relationship between these variables was significant, X^2^ (1, N = 202) = 4.728, p = 0.03. It was also unknown whether or not these family members lived with them. Most physicians (62.6%) worked more hours during the pandemic, 16.8% the same and 20.3%. less; physicians working at both Public and Private Systems had more frequently excess working hours both in virtual and personally interviews, X^2^ (4, N = 127) = 14.026, p =0.007. Also, infectologists and intensive care specialists were more likely to have excess working hours than others. X^2^ (8, N = 203) = 30.15, p < 0.001. Also, ANOVA showed that mean age of physicians who worked excess hours was lower than those who did not, F (2,200) = 3.902, p = 0.022; Post hoc comparisons using the Tukey’s B test indicated that mean age of physicians working more hours (N = 127, M = 46.89 years) was lower than those who did not (N = 42, M = 51.9 years). Almost all physicians (93%) said their stress became more intense.

Owing to stay-at-home and distancing social orders, more than half physicians stayed more hours at home, and food appeared to be the comfort of greatest choice for a high percentage (69.5%), followed by far by psychopharmaceuticals (11.9%), and alcohol intake (11.4%). A notable percentage of physicians increased some activities such as watching TV, movies, and series at home (33.2%) as well as cooking (31.7%), followed by exercising (17.8%) and reading not scientific literature (11.4%), while yoga-meditation increased in only 5.9%. Differences between men and women were found, X^2^ (4, N = 202) = 13.6, p =0.009; for instance, alcohol intake increase was higher in men than women (X^2^ (1, N = 202) = 5.55, p=0.018, while differences on other consumptions were not significant.

Considering family relationships at home, half of physicians (53.5%) said they had any change, for 25.2% were better and 21.3% considered them worse than before pandemic; more than a half answered they had no changes about loneliness, 32.7% said they were lonelier during pandemic than before, and a small percentage of physicians said they were less lonely. ANOVA showed that mean age of physicians who felt lonely during lock down was lower than those who did not, F (2,200) = 3.363, p = 0.037, Post hoc comparisons using the Tukey’s B test indicated that mean age of physicians feeling loneliness (N=67, M = 45.84 years) was lower than those who felt less loneliness (N = 14, M = 53.1 years).

Physicians expressed that difficult experiences while taking care of patients with COVID-19 were mostly related to the sadness of seeing colleagues getting very sick and isolated as well as patients arriving at their rooms assuming they will die soon and saying goodbye to their families. Also, many physicians increased their anxiety due to the disorganization at the hospitals, the lack of support of the government and hospital authorities, and the lack of the appropriate PPE as well as supplies added to the low salaries, constitute all aspects which increased their loneliness at work. Women were more likely to express distress or anxiety than men. The relationship between these variables was significant, X^2^ (1, N = 202) = 6.313, p =0.012. The frequent lack of social recognition of the extreme bad work conditions contribute to their discouragement.

In addition, many medical doctors felt very sad with daily communication of bad news to patients’ relatives as well as the long time which is required for an accurate patient evaluation. Finally, the possible own contagious, or their familiars and patients became an important cause of increased stress.

On the other hand, most physicians were not considering any career change due to their experiences treating COVID-19 patients except for considering an early retirement respect to the previously planned (41.9%). Moreover, men were more likely than women to express an anticipated retirement. The relationship between these variables was significant, X^2^ (1, N = 202) = 6.497, p =0.011.

It was interesting that the majority of physicians never closed their practices while the others reopened them very rapidly after the first pandemic days; most Medical doctors said they had a positive learning during attending COVID-19 patients; 65.1% physicians considered an obligation to treat patients with COVID-19, despite 26.1% had not the appropriate PPE and 12.9% were diagnosed with COVID-19. Notwithstanding the efforts around the world performed to develop new treatments for COVID-19, only 41.3% physicians considered them useful in their practices but 71.9% believed that, by the end of 2021, there would be a vaccine for this disease. Men compared to women considered more frequently that new drugs or vaccines would be available during 2021. The relationship between these variables was significant, X^2^ (1, N = 202) = 4.799, p =0.028.

In general, physicians did not feel an appreciation of their work during the pandemic. In fact, the majority reported distress episodes while 30% had depression and were medicated; four medical doctors had suicidal thoughts. Physicians who said to be depressed were younger than those who did not; ANOVA results showed that mean age were (F (1,201) = 3.363, p = 0.037, M = 44.73 vs M = 49.33 respectively), and for those who reported distress, statistical results were F (1,201) = 6.379, p=0.012, M = 47.15 vs M = 52.03, respectively. Most physicians (96%) considered stressful their experience during pandemic while for more than 60% was the most stressful of their practices. Considering sex differences, men expressed more frequently than women that COVID-19 patient care was the most stressing experience during their careers, X^2^ (1, N = 203) = 8.54, p = 0.003, but Kendall’s tau coefficient test found that sex is independent of ranked stress thoughts related to COVID-19 patient care (rτ = -0.75, p=0.295). Furthermore, infectologists and intensive care specialists were more likely to express that taking care of COVID-19 patients was stressful, or their most stressful experience compared with other specialties. X^2^ (12, N = 203) = 63.502, p < 0.001. Only 5 physicians reported an appreciation of their work by their authorities, while 9 by the society; relatives recognized more frequently their work (53.7%), followed by patients (24.6%), and friends (15%). Work recognition was perceived differently between men and women, X^2^ (4, N = 202) = 12.568, p = 0.014; in this sense, friends’ appreciation was higher in men than in women, X^2^ (1, N = 202) = 6.012, p= 0.014, while differences with other perceptions about work recognition were not significant. Physicians who worked at the Public System said more frequently they would choose the medical career again compared to those working at private institutions, or at both, X^2^ (2, N = 203) = 8.025, p=0.018. Nevertheless, most physicians said they would choose again the medical career.

Finally, nearly all physicians considered that the Faculty of Medicine of the National University of La Plata would bring help to their graduates to face critical and special situations, such a pandemic, and during them.

## Discussion

Physicians of La Plata have performed an extremely high effort treating COVID-19 patients in very bad conditions; they have also faced many additional, often avoidable, stressful situations. They had to deal with quick learning and training of new practices and technologies to fulfil patient care without sufficient resources and in absence of specific treatments for COVID-19, added to lack of vacations and frequent decrease of their incomes. Many medical doctors had to manage severely ill patients, care for colleagues, offer comfort to isolated dying patients, and informed family members without perceiving appreciation of the authorities and of a part of the society, which combined with a long lock down constitute a dark scenario. These facts would explain the general idea of an anticipated retirement. Also, it is interesting to highlight the initial frequent skepticism on the usefulness of new treatments reaching to the conviction of the majority that vaccines would only be available late in 2021.

Several reports showed the deleterious impact of COVID-19 pandemic on psychological and physical health of physicians and other health workers. Batra et al.^3^ performed a meta-analysis including sixty-five articles and 79.437 health workers; this study demonstrated the prevalence of anxiety (34.4%), depression (40.3%), stress (31.8%), post-traumatic stress syndrome (11.4%) insomnia (27.8%), psychological distress (46.1%) and burnout (37.4%). Chewa et al.^4^ investigated the psychological outcomes and associated physical symptoms amongst healthcare workers during COVID-19 outbreak in a multinational, multicenter study. From the 906 healthcare workers who participated in the survey, 5.3% screened positive for moderate to very-severe depression, 8.7% for moderate to extremely-severe anxiety, 2.2% for moderate to extremely-severe stress, and 3.8% for moderate to severe levels of psychological distress. Particularly, physicians showed mental disorders associated to pandemic circumstances. In fact, a study^5^ including 442 physicians showed that 64.7% had symptoms of depression, 51.6% anxiety, and 41.2% stress. This severe damage of the psychological structure of medical doctors reported is consistent with our findings. In our series, the majority of physicians (96%) considered stressful their experience during pandemic and reported distress episodes being for more than 60% the most stressful of their practices, 30% with depression and medically treated, while 32.7% felt loneliness with 4 physicians with suicidal thoughts, added to a high consumption of psychopharmaceuticals and alcohol.

Although statistically significant differences related to sex and age were found, further research would be necessary to address whether these results constitute a general finding. The small sample of the survey may be considered as a limitation of this research. At present, COVID-19 pandemic is getting worse with a record number of incidence and deaths; about 12.7% of the total population has received at least one dose of a COVID-19 vaccine, and only 1.8% fully vaccinated (https://ourworldindata.org/coronavirus/country/argentina, accessed 23 April 2021); in addition, the authorities renewed restrictions.

## Conclusions

Some authors claimed for government and healthcare agencies to protect the psychological well-being of the healthcare community, and invest resources to significantly promote the mental health of these frontline professionals^6^ as well as make efforts to reduce mental health stigma in clinical workplaces, adding “healthcare staff mental health support process”^7^. It is possible that the faculties of Medicine should contribute bringing a special milieu to help Physicians as was requested in our study. Also, WHO called team leaders and managers in health facilities to ensure that staff are aware of where and how they can access mental health and psychosocial support services and facilitate access to such services^8^.

## Data Availability

All data produced in the present work are contained in the manuscript

**All the authors have seen and approved the manuscript**.

## Statement On Ethical Approval

The research was approved by the Medical Bioethics Committee, Faculty of Medical Sciences, National University of La Plata, Argentina, reference N°101/21.

## References

1. Mehta S, Machado F, Kwizera A, et al. COVID19: a heavy toll on health-care workers. The Lancet 2021; 9: 226–228 doi:https://doi.org/10.1016/S2213-2600(21)00068-0).

2. Medscape US and International Physicians’ COVID-19 Experience Report: Risk, Burnout, Loneliness (https://www.medscape.com/slideshow/2020-physician-covid-experience-6013151) Leslie Kane, MA | September 11, 2020.

3. Batra K, Pal Singh T, Sharma M, Batra R, Shevaneveldt N. The Impact of COVID-19 on Healthcare Worker Wellness: A Scoping Review Investigating the Psychological Impact of COVID-19 among Healthcare Workers: A Meta-Analysis. Int J Environ Res Public Health 2020; 17: 1–33; 2020. https://www.ncbi.nlm.nih.gov/pmc/articles/PMC7730003/

4. Chewa NWS, Leeb GKH, Tanb BYQ, et al. A multinational, multicentre study on the psychological outcomes and associated physical symptoms amongst healthcare workers during COVID19 outbreak. Brain, Behavior, and Immunity 2020; 88: 559–565. https://www.ncbi.nlm.nih.gov/pmc/articles/PMC7172854/

5. Elbay RY, Kurtulmuş A, Arpacioğlu S, Karadere E. Depression, anxiety, stress levels of physicians and associated factors in Covid-19 pandemics. Psychiatry Research 2020; 290: 1–5.

6. Ornell F, Chwartzmann Halpern S, Paim Kessler FH, Correa de Magalhaes Narvaez J. The impact of the COVID-19 pandemic on the mental health of healthcare professionals. Cad. Saúde Pública 2020; 36 :1–6. https://doi.org/10.1590/0102-311x00063520

7. Galbraith N, Boyda D, McFeeters D, Hassan T. The mental health of doctors during the Covid-19 pandemic. B J Psych Bull. 2020 Apr 28 : 1–4. https://www.ncbi.nlm.nih.gov/pmc/articles/PMC7322151/

8. World Health Organization. Mental health and psychosocial considerations during the COVID-19 outbreak. https://www.who.int/publications/i/item/WHO-2019-nCoV-MentalHealth-2020.1

